# Understanding cerebral blood flow dynamics for Alzheimer’s disease prevention through acute exercise (flADex): Protocol for a randomized crossover trial

**DOI:** 10.1101/2024.12.28.24319064

**Authors:** Isabel Martín-Fuentes, Beatriz Fernandez-Gamez, Sol Vidal-Almela, Alfredo Caro-Rus, Patricio Solis-Urra, Lucía Sánchez-Aranda, Javier Fernández-Ortega, Javier Sanchez-Martinez, Andrea Coca-Pulido, Marcos Olvera-Rojas, Emilio J. Barranco-Moreno, Jose D. Marin-Alvarez, Esmée A. Bakker, Angel Toval, Darío Bellón, Alessandro Sclafani, Thomas K. Karikari, Kirk I. Erickson, Manuel Gómez-Río, Francisco B. Ortega, Irene Esteban-Cornejo

## Abstract

**INTRODUCTION:** Alzheimer’s disease (AD) is a leading cause of disability worldwide. Alterations in cerebral blood flow (CBF) and AD blood biomarkers are fundamental at early stages of AD. Exercise shows promise in delaying physiological changes, but its mechanisms for enhancing brain health remains unclear. FlADex aims to examine the acute effects of different exercise types on CBF and blood biomarkers in older adults. This protocol describes the methodology and rationale of flADex.

**METHODS:** FlADex is a counterbalanced crossover trial that will include 20 older adults aged 68 to 83 years old with negative brain amyloid status (<12 centiloid) and APOEε4 noncarriers. Participants will complete a 30-minute session of each condition in a randomized order: (i) moderate-intensity aerobic exercise (60-70% maximal heart rate), (ii) moderate-intensity resistance exercise (rating of perceived exertion: 4-6 points out of 10), and (iii) resting condition. Changes in the CBF is the primary outcome and will be assessed by magnetic resonance imaging using pseudo-continuous arterial spin labeling at pre- and at 3 timepoints post-condition (starting at 20, 27, 34 minutes). Secondary outcomes are biomarkers of AD pathology and neurodegeneration (Aβ42, Aβ40, p-tau217, p-tau181, BD-tau, GFAP, NfL) and growth factors (BDNF, IGF-1), measured through blood samples collected at pre- and post-condition (at 3, 50, 70 minutes). Moreover, cognitive outcomes and mood status will be measured pre- and post-condition.

**DISCUSSION:** FlADex will highlight the acute effects of exercise types on CBF and biomarkers before beta-amyloid accumulation. Acute effects on CBF dynamics and blood biomarkers are expected to be greater with aerobic than resistance exercise when compared to resting. CBF is expected to vary by brain region, and biomarkers to fluctuate dynamically post-exercise. This will provide critical insights into exercise impact on vascular and molecular pathways associated with AD pathology, and potential recommendations for standardized blood sampling to enhance diagnostic accuracy.

**Highlights:**

- Exercise may delay the development of AD, but its physiological mechanisms are unclear.
- flADex explores exercise-induced changes in CBF and blood biomarkers.
- 20 older adults will undergo a bout of aerobic, resistance exercise, and resting.
- CBF may increase more after aerobic exercise, with changes varying by brain region.
- Standardized rest conditions before blood sampling may enhance diagnostic accuracy.

**Declarations of interest:** TKK has consulted for Quanterix Corp., has received honoraria from the NIH for study section membership, and honoraria for speaker/grant review engagements from UPENN, UW-Madison, Advent Health, Brain Health conference, Barcelona-Pittsburgh conference and CQDM Canada, all outside of the submitted work. TKK has received blood biomarker data on defined research cohorts from Janssen and Alamar Biosciences for independent analysis and publication, with no financial incentive and/or research funding included. TKK is an inventor on patent #*WO2020193500A1* and patent applications #*2450702-2, #63/693,956, #*63/679,361, and *63/672,952*.

## 1. BACKGROUND

Alzheimer’s Disease (AD) is a leading cause of disability and dependency worldwide [1], characterized by a gradual deterioration in cognitive and functional abilities. Pathophysiological signs of AD begin approximately10 to 20 years before the onset of symptoms of cognitive decline [2,3]. The accumulation of amyloid-beta (Aβ) plaques and neurofibrillary tangles composed of hyperphosphorylated tau protein (p-tau) are well-established pathological hallmarks of AD. Additionally, cerebral blood flow (CBF) has gained increasing attention as a key vascular pathway in AD research. Indeed, CBF dysregulation can be observed in older adults at risk for AD, even before the appearance of Aβ accumulation or brain atrophy [2,4]. Disrupted CBF exacerbates neurodegeneration by upregulating the BACE1 enzyme –which produces Aβ – and by promoting the accumulation of p-tau. This reduction in CBF may significantly contribute to cognitive decline by either triggering the amyloid cascade or amplifying Aβ production through a feedback loop [5].

The chronic neuroprotective benefits of exercise likely result from the repeated acute effects that occur during or following each exercise bout, as bodily systems respond to meet the increased metabolic demands [6,7]. Observational and animal studies suggest that chronic exercise is associated with increased CBF and angiogenesis in the brain [6,8]. During aerobic exercise, CBF often follows an inverted U response, increasing at the start of exercise, plateauing, and then declining towards the end of exercise [9,10]. However, after exercise, the cerebrovascular effects vary across brain regions and over time, depending on the type of exercise (e.g., aerobic vs. resistance) [10–12]. Further, emerging evidence suggests that exercise may acutely alter the concentrations of neurodegenerative biomarkers (e.g., Aβ, neurofilament-light chain [NfL]) and growth factors (e.g., brain-derived neurotrophic factor [BDNF], insulin-like growth factor 1 [IGF-1]) in the bloodstream [13,14]. These changes are hypothesized to influence cognitive function, potentially mediating the brain health benefits of exercise [7,8]. However, the acute implications of exercise-induced CBF changes on blood biomarkers and cognitive function, following different types of exercise, remain underexplored. Investigating this is also crucial for establishing recommendations on whether blood samples for biomarkers of AD pathology and neurodegeneration should be collected under standardized rest conditions prior to measurements.

Research on acute responses to exercise, using neuroimaging combined with measures of CBF, blood biomarkers, and cognitive function, is essential to reveal how exercise enhances brain health and aids in preventing or managing AD [13]. Therefore, flADex will examine the acute effects of aerobic and resistance exercise compared to a resting condition on CBF, blood biomarkers, and their cognitive implications in older adults. This protocol describes the rationale and methodology of the flADex trial.

### Trial objectives

The *primary objective* is to examine the acute effects of a bout of aerobic exercise, resistance exercise and a resting condition on global and regional CBF using cutting-edge magnetic resonance imaging (MRI) in older adults.

The *secondary objectives* are to examine: (i) the acute effects of a bout of aerobic exercise, resistance exercise and a resting condition on blood biomarkers of AD pathology and neurodegeneration (Aβ42, Aβ40, p-tau217, p-tau181, brain-derived tau [BD-tau], glial fibrillary acidic protein [GFAP] and NfL) and growth factors (BDNF and IGF-1); (ii) the acute effects of a bout of aerobic exercise, resistance exercise and a resting condition on cognitive and mood outcomes; (iii) whether exercise-induced changes in CBF mediate changes in blood biomarkers (or vice versa); and (iv) whether exercise-induced changes in CBF or blood biomarkers mediate changes in cognitive and mood outcomes.

Our *overall hypothesis* is that aerobic exercise will lead to a greater increase in regional CBF compared to resistance exercise [7,15] particularly when contrasted with resting, and may also differentially impact blood biomarkers [16–18].

## 2. METHODS

### 2.1 Trial design and ethics

The flADex trial will follow a within-subject crossover design with pre- and post-test assessments in 20 individuals (balanced sex distribution) aged 68-83 years old. Each participant will be included in three experimental conditions (i.e., aerobic exercise, resistance exercise and resting) in a randomized order (n=3 x 20 observations) (see **Figure 1**). The trial will be performed in two research institutes at the University of Granada (Spain): (i) Sports and Health University Research Institute (iMUDS) for the familiarization visit; and (ii) Mind, Brain and Behaviour Research Centre (CIMCYC) for the experimental condition visits.

**Figure 1.**
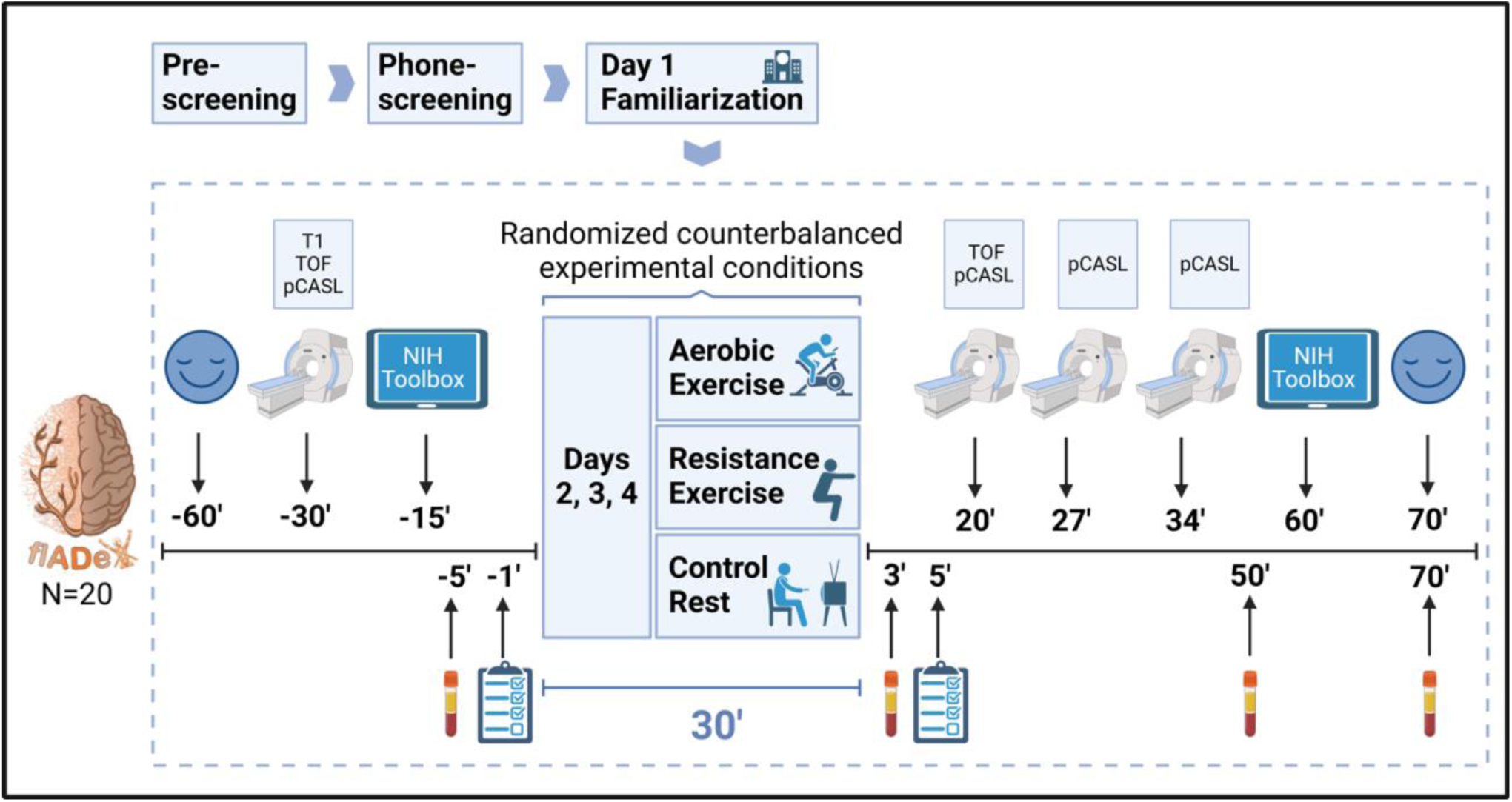
Trial design and key measurement timepoints of the flADex trial. MRI: 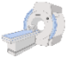; Episodic memory and inhibition/attention: 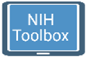; AD and neurodegenerative blood biomarkers, and growth factors: 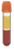; Mood (POMS): 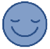; Enjoyment (PACES) and feelings of (dis)pleasure (Feeling Scale): 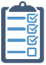 T1: T1-weighted MPRAGE structural; TOF: Time of flight angiography; pCASL: Pseudo Continuous Arterial Spin Labeling.

The trial, which has been registered in ClinicalTrials.gov (NCT06584656; approval: 04/09/2024), will follow the principles of the Declaration of Helsinki, and has been approved by the Research Ethics Board of the Andalusian Health Service (CEIM/CEI Provincial de Granada; #SICEIA-2024-000602; approval: 30/04/2024). All participants will provide written informed consent. Procedure manuals of the flADex trial will be made available to allow other researchers to replicate the results (https://github.com/fladexprojectugr). flADex has been designed following the Standard Protocol Items for Randomized Interventional Trials (SPIRIT) [19], the SPIRIT-Outcomes 2022 Extension [20] (Supplementary Table A1), and is reported in line with the Consolidated Standards of Reporting Trials (CONSORT) 2020 statement for extension to randomized crossover trials [21]. Any substantial modifications to the protocol will be communicated to the trial registry, detailed in the main results paper, and submitted for review to the Research Ethics Board.

### 2.2 Eligibility criteria, screening of participants and recruitment

The flADex trial will recruit individuals from the previous AGUEDA (Active Gains in brain Using Exercise During Aging) trial [22,23]. Recruitment and screening are illustrated in **Figure 2**. Of the 53 eligible individuals from AGUEDA, 20 will be eventually included and randomized for the crossover trial (balanced sex distribution).

**Figure 2.**
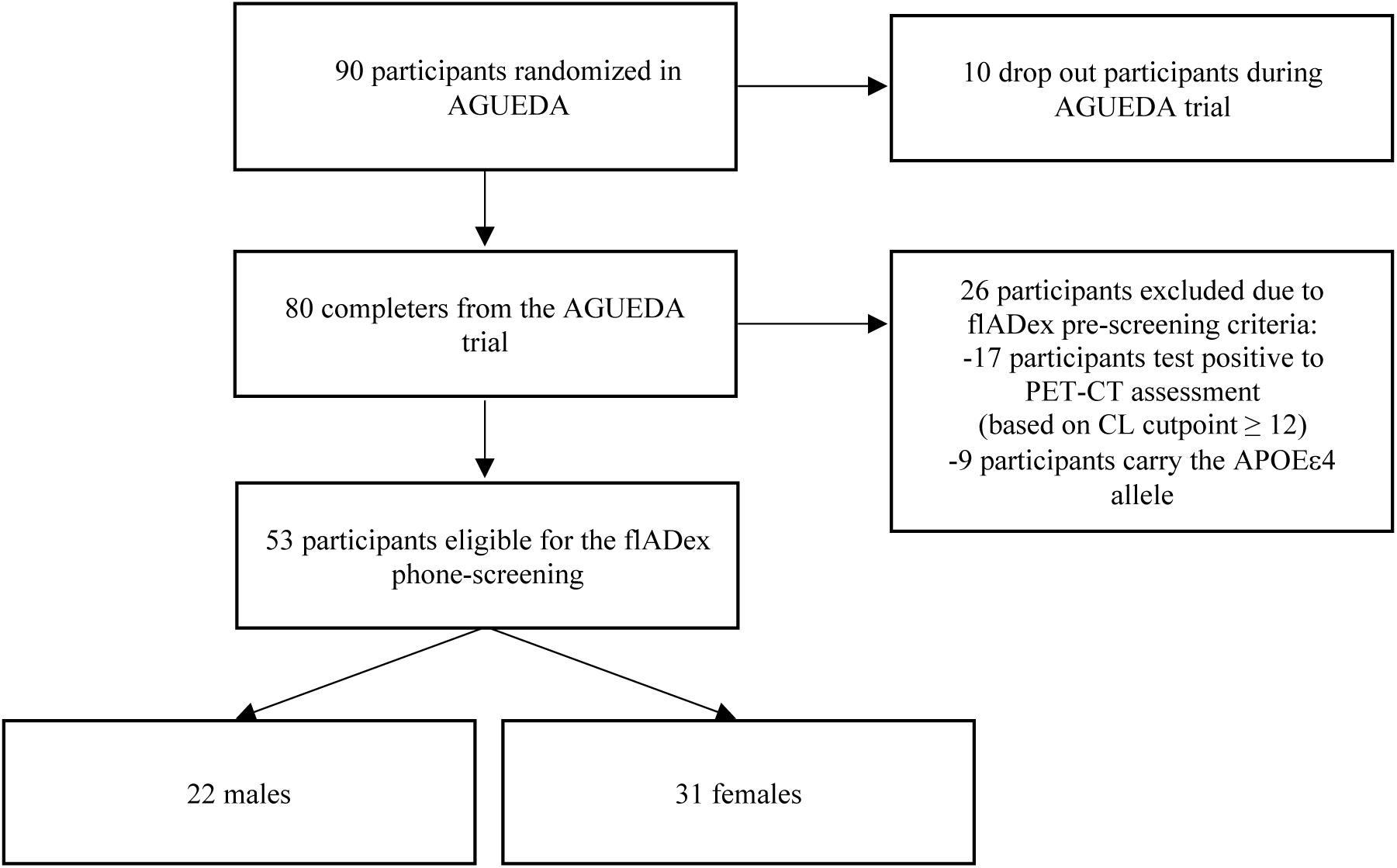
Recruitment and screening of participants for the flADex trial. PET-CT, Positron Emission Tomography - Computed Tomography; CL, centiloid; APOEε4: Apolipoprotein E 4 allele.

Eligibility criteria assessed through pre-screening and phone screening are detailed in **Table 1**. During the pre-screening phase, inclusion criteria will be checked using the AGUEDA database. Eligible participants will undergo phone screening, and if no exclusion criteria are identified, they will be scheduled for a familiarization visit (day 1).This visit includes signing informed consent; administering the Montreal Cognitive Assessment test (MoCA) [24]; completing questionnaires related to MRI compatibility, medical information, blood baseline conditions and physical activity; measuring weight and height (to calculate body mass index) and resting heart rate; and explaining and practicing exercise conditions. Participants who successfully complete the familiarization visit will be included and randomized (days 2,3,4), with recruitment set to begin in September 2024.

**Table 1.** Eligibility criteria of the flADex trial.

| Inclusion Criteria<sup>†</sup> | Exclusion Criteria<sup>‡</sup> |
| --- | --- |
| Adults aged between 68 and 83 years old who participated in the AGUEDA trial.* | Ambulatory with pain or regular use of an assisted walking device. |
| Non-pathological cerebral beta-amyloid status (based on a Centiloid cut-point <12, measured by PET-CT). | Not living in community settings during the trial. |
| APOEε4 negative status. | Pathological diagnosis related to any physical or mental condition that would preclude the participant from exercising or performing the tests. |
|  | MRI incompatibility. |

<sup>†</sup>Pre-screening criteria; <sup>‡</sup>Phone-screening criteria; \*Participants from the AGUEDA trial were 65 to 80 years old at enrolment and are now 3 years older, making their age range 68 to 83 years for flADex.

### 2.3 Randomization

Each participant will be randomized after familiarization to all trial conditions: (A) bout of aerobic exercise, (B) bout of resistance exercise, and (C) resting condition. The order of each condition will be randomized for each participant to achieve a counterbalanced crossover design. A 3x3 Latin square design has been implemented for randomization [25], utilizing the “rlatin” function from the “magic” package in R Studio. A randomization dashboard has been created (https://ersanchez3.shinyapps.io/latin_square_design/) and will be executed for each wave of participants by a blinded researcher external to the trial (Dr. Cabanas-Sanchez, Autonomous University of Madrid, Spain). To prevent selection bias, allocation concealment will keep the assignment sequence hidden from those enrolling participants, ensuring random and unbiased order assignments.

### 2.4 Blinding

Participants will inherently become aware of their assigned condition at the start of each visit. However, the principal investigator, MRI technician, and research staff responsible for processing MRI data and conducting statistical analyses will remain blinded to the participants’ condition for each day. Blinding ensures results are free from researcher bias or expectations.

### 2.5 Overview of experimental conditions

Each participant’s trial will last three weeks, preceded by the familiarization visit. Participants will complete three 30-minute experimental conditions, each separated by a one-week washout period, with a ratio of one trainer per participant. The aerobic and resistance exercise bouts will include a 4-minute warm-up and a 26-minute exercise session (see **Figure 3**). All conditions will be performed after 12 hours fasting with a standardized dinner the previous night.

**Figure 3.**
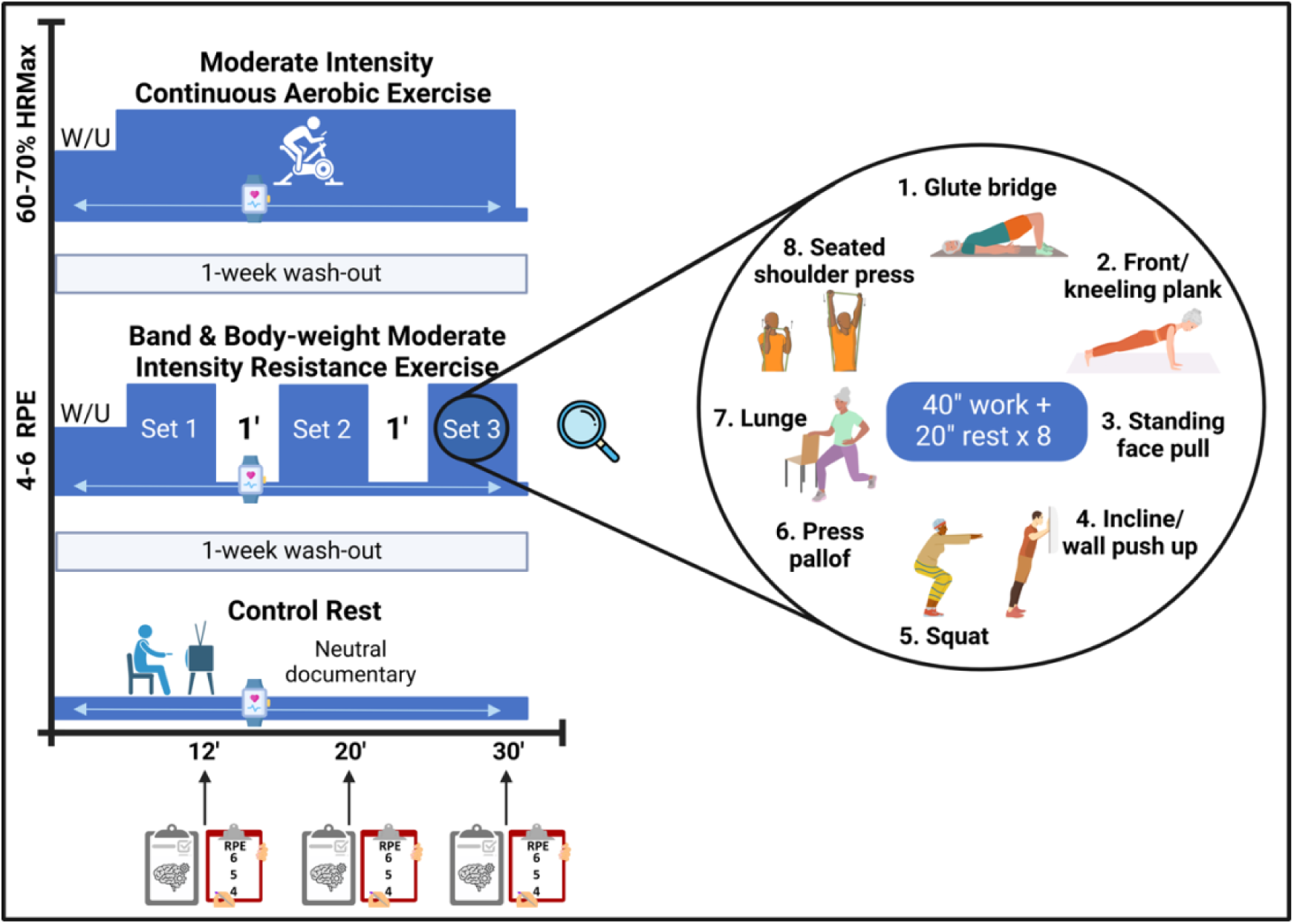
Experimental conditions of the flADex trial. HRMax: Maximum heart rate; RPE: Rating of perceived exertion; W/U: Warm-up.

#### 2.5.1 Aerobic exercise condition

Participants will perform continuous moderate-intensity aerobic exercise on a cycle ergometer (Keyser m3) at 60%–70% of their maximal heart rate (HRmax; calculated as 220 – age) [32]. To achieve the target heart rate, the intensity will be adjusted according to the participant’s response, with a primary focus on increasing the ergometer’s resistance rather than the cadence. For further details, refer to Supplementary Material A.2.2.

#### 2.5.2 Resistance exercise condition

Participants will perform a combination of upper- and lower-body exercises using elastic bands with different resistance levels and their body weight as primary resistance. The target intensity will be based on 4-6 points of rate of perceived exertion using the OMNI-Resistance Exercise Scale (RPE, 0– 10) [26]. Participants will perform three sets of eight different exercises, with each exercise lasting 40 seconds followed by 20 seconds of rest and a 1-minute rest period between sets. Exercises might be adapted accordingly to the participants’ ability or situation (e.g., blood line placement). The exercises will include glute bridge, front plank, standing face pull, incline push-up, squat, press pallof, walking lunge and seated shoulder press. See supplementary material A.2.1 for extended information.

#### 2.5.3 Resting condition

Participants will remain seated and isolated in a room, watching a standardized neutral documentary on a tablet for 30 minutes without any cognitive engagement. (https://www.youtube.com/watch?v=WLeTofB2wbo&t=1531s).

### 2.6 Outcomes

**Table 2** summarizes the primary and secondary outcomes, and exercise-related variables, along with the assessment instruments and data collection time points.

**Table 2.** Outcomes, instruments and time points of data collection of the flADex trial.

| Primary outcomes | Before condition | During condition | After condition | Instrument / specific outcome / unit |
| --- | --- | --- | --- | --- |
| **Cerebral Blood Flow** | -30' | - | 20', 27' and 34' | MRI: turbo gradient spin echo-pseudo continuous arterial spin labeling (pCASL)<br>mL/100 g/min |
| Secondary outcomes | Before the condition | During the condition | After the condition | Instrument / specific outcome / unit |
| **Mood status** | -60' | - | 70' | Scale Profile of Mood States (POMS)<br>15-items scale<br>Total mood disturbance: 90-144 total score |
| **Episodic memory** | -15' | - | 60' | NIH Toolbox:<br>Picture Sequence Memory Test<br>Raw score n total correct items |
| **Inhibition/attention** | -15' | - | 60' | NIH Toolbox:<br>Flanker test<br>Incongruent trials inverse efficiency score |
| **Blood biomarkers of AD pathology and neurodegeneration** | -5' | - | 3', 50' and 70' | Plasma samples<br>Aβ42/40 ratio, p-tau217, p-tau181, BD-tau, GFAP and NfL<br>pg/mL |
| **Growth factors** | -5' | - | 3', 50' and 70' | Serum samples<br>BDNF, IGF-1<br>pg/mL (BDNF), ng/mL (IGF-1) |
| **Emotional response** | -1' | - | 1' | Feeling scale<br>11-points scale<br>-5 to 5 total score |
| Exercise-related variables | Before condition | During condition | After condition | Instrument / specific outcome / unit |
| **Rate of Perceived Exertion (only aerobic and resistance condition)** | - | 12', 20' and 30' | - | OMNI-Resistance Exercise Scale (OMNI-RES)<br>10-points scale<br>1-10 total score |
| **Cognitive engagement** | - | 12', 20' and 30' | - | Cognitive Load Measurement scale<br>9-points scale<br>1-9 total score |
| **Heart Rate** | - | During 30' | - | Polar H10<br>Heart Rate per second<br>beats/sec |
| **Repetitions in reserve (only resistance condition)** | - | After each exercise | - | The Repetitions in Reserve (RIR)<br>Number of repetitions<br>n |
| **Enjoyment** | - |  | 5' | Physical activity Enjoyment scale (PACES)<br>8-items scale<br>7-56 total score |
| **Blood pressure** | -60' |  | 1' | Omron M3<br>Systolic and diastolic blood pressure<br>millimeters of mercury (mmHg) |
| **Temperature** | -50' |  | 20' | Digital Infrared Forehead Thermometer (iPiccoli)<br>Temperature<br>degrees Celsius (°C) |

MRI, Magnetic Resonance Imaging; Aβ42/40, ratio Amyloid-beta 42/ Amyloid-beta 40; p-tau217, p-tau181, phosphorylated tau protein at positions 217 and 181; BD-tau, Brain-Derived tau; GFAP, Glial Fibrillary Acidic Protein; NfL, Neurofilament Light Chain; BDNF, Brain-Derived Neurotrophic Factor; IGF-1, Insulin-Like Growth Factor 1.

#### 2.6.1 Primary outcomes

##### Change in CBF

CBF will be assessed by MRI, using a Siemens Magnetom PRISMA Fit 3T scanner with a 64-channel head coil located at CIMCYC. Specific acquisition parameters for MRI are detailed in **Table 3**. Body temperature and any incidental bodily conditions will be registered before MRI acquisitions to ensure physiological stability. pseudo-continuous Arterial Spin Labeling (pCASL) sequence are used to determinate global and regional CBF in resting supine position condition. A structural T1 sequence (only pre-condition) will be used to co-register the pCASL and delineate regions of interest for CBF. Time-of-flight angiography (TOF) (before pCASL pre-condition and before first pCASL post-condition) sequence will be used to identify the carotid arteries. The preprocessing analysis of pCASL will be carried out using the most up-to-date pipelines (ASLprep).

**Table 3.** MRI parameters for the flADex trial.

| Sequence | Parameters | Measure | Acquisition time (min) |
| --- | --- | --- | --- |
| TGSE pCASL | Transversal, resolution = 2.5x2.5x3.0mm, TR = 4100ms, TE = 36.8 ms, Background suppression, M0 and 12 label-control pairs, Postlabeling delays = [500, 500, 1000, 1000, 1500, 2000, 2000, 2000, 2500, 2500, 2500 ms], Labeling duration = 1500 ms, 4 segment readout | Cerebral blood flow | 7 |
| T1-weighted MPRAGE structural | Sagittal, 0.8 mm isotropic resolution, TR = 2400 ms, TE = 2.31 ms, TI = 1060 ms, FOV = 256 mm, slices = 224 | Brain structure | 6 |
| TOF (Time of flight angiography) | Resolution = 0.8x0.8x1.2 mm3, TR = 19 ms, TE = 2.82 ms, Flip angle = 18grad, FOV = 200 mm, acquisition bandwidth = 219 Hz/Px, slices = 32 (*oversampling 25%*) | Angiography (cerebral vascularization) | 1 |

TGSE pCASL, Turbo Gradient Spin Echo Pseudo-Continuous Arterial Spin Labeling; TR, Repetition Time; TE, Echo Time; TI, Inversion Time; FOV, Field of View; MPRAGE, Magnetization Prepared Rapid Gradient Echo.

#### 2.6.2 Secondary outcomes

##### Change in blood biomarkers of AD pathology and neurodegeneration, and growth factors

Aβ42, Aβ40, p-tau217, p-tau181, BD-tau, GFAP, NfL, BDNF and IGF-1 will be assayed. Blood samples will be collected at CIMCYC facilities by a qualified nurse. Each day will involve four draws with one puncture: one pre-condition (using a bloodline) followed by draws at 3, 50, and 70 minutes after the condition. During each session, 48 ml of blood (3 tubes –2 for plasma and 1 for serum–, * 4 ml/each * 4 draws) will be collected. Samples will be kept cold during collection, processed according to regulations [27], pseudoanonymized, and stored at −80°C at iMUDS for analyses after trial completion. Blood biomarkers of AD pathology and neurodegeneration will be measured using single molecular array (Simoa) following manufacturer’s instructions at the University of Pittsburgh [28].

##### Change in episodic memory

The Picture Sequence Memory Test from the Cognitive NIH Toolbox (a computer-based battery which has been validated in Spanish) measures episodic memory. The raw score from the cumulative number of adjacent pairs of pictures remembered correctly over two learning trials will be used as outcome [29].

##### Change in inhibition/attention

The Flanker test from the Cognitive NIH Toolbox measures inhibitory control and attention. The outcome will be the inverse efficiency score of incongruent trials calculated as reaction time/accuracy (RT/ACC) [29].

##### Mood status

A shortened version of the Profile of Mood States (POMS) scale will be used to assess mood status [30]. It consists of adjectives or mood descriptors, where individuals rate how they have been feeling on a scale typically ranging from “Not at all” (0) to “Extremely” (4). The shortend version includes 15 items divided into 5 dimensions: depression, vigor, anger, tension, and fatigue. The final score, calculated as: [depression]+[anger]+[tension]+[fatigue] – [vigor], reflects total mood disturbance, with higher scores indicating more negative emotions and less positive energy.

##### Emotional response

The feeling scale [31] will be used to assess emotional response. It is an 11-point scale ranging from -5 (very bad) to +5 (very good) used to measure an individual’s feelings in terms of pleasure or displeasure.

#### 2.6.3 Exercise-related variables

##### Rate of Perceived Exertion (RPE)

RPE will be assessed using the OMNI-Resistance Exercise Scale (OMNI-RES) of perceived exertion from 0-10 points [26].

##### Cognitive engagement

The cognitive load scale is a 9-item scale used to assess the mental effort or cognitive load experienced by individuals when engaging in a task, particularly in educational or learning contexts. It involves a self-reported measure where participants rate their perceived mental effort on a scale, ranging from 1-very low mental effort to 9-very high mental effort [32,33].

##### Heart rate (HR)

HR will be continuously monitored second-by-second during all three conditions using a Polar H10 monitor, which includes a chest strap and wristwatch.

##### Repetitions in reserve (RIR)

RIR is a self-regulation technique used in resistance training to gauge exercise intensity. It involves estimating the number of additional repetitions a participant could perform before reaching muscle failure after completing a set [34].

##### Enjoyment

Enjoyment of physical activity will be assessed using the reduced and validated version of the physical activity enjoyment scale (PACES) [35], an 8-item scale assessing in a range of 1 to 7 a series of sensations or moods with respect to its opposite.

##### Blood pressure (BP)

BP will be measured using a validated automated monitor (Omron M3, Intellisense, OMRON Healthcare Europe, Spain) with participants seated and their left arm at heart level. After 5 minutes of rest, two readings will be taken at 1-minute intervals, and the average of the readings will be used for analysis.

##### Temperature

Body temperature will be measured using a digital infrared forehead thermometer (iPiccoli, AET-R1B1), with participants standing, ensuring their forehead is clean and free from sweat.

### 2.7 Safety and adverse events

The expected risks and side effects from participating in the flADex trial are low [18,36,37]. Any unusual symptoms (e.g., dizziness, chest pain) or adverse events during the trial will be noted by the research staff. Participants may withdraw from the trial at any time, and the research staff may terminate a participant’s involvement if any minimal risk to their safety is identified.

During exercise conditions, participants may experience very mild symptoms/sensations: *(i) Expected physiological symptoms/sensations:* These include fatigue, muscle soreness, shortness of breath, and rapid heart rate. Especially for those not accustomed to riding a cycle ergometer, muscle soreness may be felt after a session of exercise, and *(ii) Unexpected adverse symptoms/events that could be triggered by physical activity:* These include irregular heart rate, chest pain, headache, nausea, dizziness, muscle cramps, musculoskeletal injuries, vomiting, intense chest pain, malignant arrhythmia, or cardiac arrest. However, the experimental conditions and tests will be conducted in a medically supervised environment equipped to handle emergencies, with a team prepared to address any adverse event.

Additionally, minor adverse effects may be experienced in specific assessments; *(i) MRI:* Participants might experience mild claustrophobia; however, trained technical and trial staff will be available to provide support as needed, and *(ii) blood draws:* potential effects include bruising, dizziness, skin irritation, swelling, or pain, yet these will be minimized through the expertise of the healthcare staff collecting the blood. Adverse events will be categorized based on predefined criteria using common terminology criteria for adverse events as mild, moderate and severe [18].

### 2.8 Sample size

Previous studies on acute exercise have shown that a single bout of exercise changed CBF by 15% [10] and 9% [11]. Specifically, the sample size is based on the mean changes (M) in CBF of 6 ml/100g/min (M1=40.5 and M2=34.2) with a standard deviation (SD) of 6.46 ml/100g/min [3]. Considering the calculation of Cohen’s d based on previous studies (d=M1-M2/combined SD), a large effect size (Cohen’s d = 0.9) is expected [3]. Therefore, using an alpha of 0.05 and a standard power of 80%, a sample size of 20 is needed. Participants who withdraw from the trial will be replaced to ensure sufficient power to assess our primary outcome. Thus, the experiment will be completed when 20 participants have completed all three experimental conditions. If a participant misses one visit, it will be rescheduled within 2-weeks of the last completed visit. This sample size is feasible based on our previous experiences involving MRI and exercise interventions.

### 2.9 Data analysis plan

#### 2.9.1 Brief description of the primary and secondary analyses

##### Effects on primary and secondary outcomes

The primary and secondary outcomes will be analyzed following a per-protocol approach using linear mixed model. The model will include fixed effects for time (four or two levels depending on the outcome), condition (three levels), time*condition interaction, as well as the unique participant identifier as a random effect. The number and patterns of missing data will be explored and reported. Missing data will be assumed to be missing at random and handled within the linear mixed model analyses. Model assumptions will be assessed. If violations are detected, appropriate corrective measures will be implemented, including data transformations. Condition effects will be evaluated using a time-by-treatment interaction term and described with estimated marginal means and 95% confidence intervals. All statistical tests will be two-tailed. *P* for significance will be set at 0.05.

##### Mediation and correlation analysis

Mediation analysis will be performed following AGReMA (A Guideline for Reporting Mediation Analyses) recommendations [43], using CBF and blood biomarkers as mediators depending on the outcome. When blood biomarkers are the outcomes, CBF will act as the mediator, and vice versa. When cognitive and mood indicators are the outcomes, both CBF and blood biomarkers will act as mediators. In addition, bivariate correlations will be performed between changes in CBF, changes in blood biomarkers and changes in cognitive and mood outcomes.

##### Exercise intervention parameters

Parameters related to the exercise conditions (i.e., RPE, cognitive engagement, HR, RIR and enjoyment) will be presented in a descriptive manner. These parameters will help, for instance, to determine whether participants reached the target intensity levels.

#### 2.9.2 Exploratory analyses

Moderation analysis will be performed using the experimental condition as the independent variable (aerobic vs resistance vs resting), CBF as outcome, and sex as moderator. Results will be interpreted cautiously due to the limited sample size and potential lack of statistical power for moderation effects. Nevertheless, performing this analysis remains informative for understanding exploratory trends in how sex may moderate acute responses to different exercise types.

### 2.10 Data management and sharing

The flADex trial will use two systems for data storage: the REDCap platform for secure, traceable handling of non-imaging data and a dedicated desktop computer for pseudonymized participant data and MRI scans. Physical forms will be securely stored at the iMUDS facility, with sensitive data replaced by artificial identifiers and the key linking identities stored separately. Quality control includes imaging data inspection and REDCap validation. The protocol and analysis plan are registered on ClinicalTrials.gov, with metadata made publicly available under FAIR principles and records assigned DOIs. Results will be shared through journals, conferences, and public platforms, following ethical and governance standards. Reproducibility is promoted through open sharing of protocols, analysis, and data management plans. Pseudonymized data will be available under restricted access 12 months post-publication, adhering to EU-GDPR and participant consent. All data produced in the present study will be available upon reasonable request to the authors. No data monitoring board is required under Spain’s funding rules.

## 3. DISCUSSION

The flADex trial seeks to advance the understanding on the acute effects of different exercise modalities on CBF, blood biomarkers, and cognitive function in older adults. The findings may reveal mechanisms through which exercise delays or slows cognitive decline, particularly vascular and molecular pathways associated with AD pathology in aging populations at risk for the disease.

flADex is expected to reveal distinct CBF response patterns between aerobic and resistance exercise. A systematic review (N=52 studies) showed that during a bout of aerobic exercise, CBF generally follows an inverted U-shaped response [9]. This autoregulatory mechanism ensures the increased oxygen and nutrient demands during exercise are met, while preventing vascular overloading, which is particularly relevant in aging populations prone to cerebrovascular dysregulation [38,39]. Additionally, acute increases in CBF, particularly in the hippocampus—a critical region for memory—, have been observed following aerobic exercise [39], and 15, 40, and 60 min after exercise cessation [12] suggesting enhanced vascular plasticity and a potential protective effect against cognitive decline. However, evidence on acute CBF changes following other types of exercise (e.g., resistance training) and across different brain regions remains limited, with fluctuations expected to vary depending on the brain region [39], highlighting the need for further investigation.

Chronic aerobic and resistance exercise have complementary effects on brain health. Aerobic exercise supports gray matter preservation, reduces systemic inflammation, and improves lipid profiles [40,41], while resistance training benefits white matter integrity and influences muscle-related biomarkers like creatine kinase and anabolic hormones (e.g., testosterone and IGF-1) [36,42]. Acutely, both exercise modalities induce inflammatory and oxidative stress; however, aerobic exercise more effectively improves insulin sensitivity and lipid metabolism, while resistance training elicits greater inflammation due to muscle damage [7,22,38]. flADex will provide novel insights into the distinct and overlapping mechanisms through which exercise acutely impacts CBF and its cognitive implications in older adults. Understanding these acute effects could help optimize chronic exercise interventions tailored to individual cerebrovascular and metabolic needs.

Beyond CBF, flADex will measure changes in blood biomarkers of AD pathology and neurodegeneration and growth factors which are critical to understanding AD and brain health. For instance, emerging evidence suggests that aerobic exercise may acutely reduce levels of NfL, a marker of axonal damage and neuronal injury [14], and modulate the kynurenine pathway, enhancing the production of neuroprotective metabolites [43]. Similarly, resistance exercise, despite inducing greater oxidative stress and inflammation due to muscle repair processes, might also promote adaptive mechanisms that contribute to neuroprotection [14]. BDNF and IGF-1 are also vital for brain health by supporting neuronal survival and synaptic growth and aiding brain repair and cognitive function, respectively. Acute aerobic and resistance exercise significantly increase serum BDNF and IGF-1 concentrations post-exercise in older men (60 mean age) compared to controls [17]. Then, measuring these biomarkers highlights how acute exercise-induced changes impact neuronal integrity, inflammation, and cognition, advancing on the molecular pathways of exercise for AD prevention and management during aging. Additionally, examining the acute effects of exercise on biomarkers of AD and neurodegeneration will help determine whether blood biomarker sampling should be performed under standardized rest conditions prior to measurements to improve diagnostic accuracy.

The flADex trial has a few limitations; while the sample size (20 participants) was calculated to test the primary hypothesis, some secondary analyses may be underpowered, which could limit their statistical reliability and generalizability. Additionally, unmeasured factors like daily activities or diet may influence outcomes despite the counterbalanced crossover design minimizing confounders; however, implementing standardized conditions (e.g., evening meal) before each experimental day will minimize these potential influences. Notable strengths of this trial include its innovative crossover design, which reduces variability by using participants as their own controls and maximizes statistical efficiency in a trial with small sample. Advanced imaging and biomarker analyses provide detailed insights into the acute effects of exercise on CBF and blood biomarkers. Additionally, the identification of acute CBF fluctuation patterns in different brain regions during exercise represents a significant contribution to understanding exercise-induced neurovascular responses. Another key strength lies in the association of these CBF fluctuations with blood biomarkers and cognitive function, offering a comprehensive perspective on the interplay between the underlying mechanisms. Furthermore, comparing aerobic and resistance exercise highlights distinct impacts, while the focus on preclinical AD in a specific subgroup targets an optimal stage for delaying symptom progression. Lastly, this study is critical for establishing recommendations on whether blood samples for biomarkers of AD pathology and neurodegeneration should be collected under standardized rest conditions prior to measurements, similar to other physiological factors such as food intake, particularly when using these biomarkers for diagnosis, monitoring disease progression and evaluating drug responses [44].

In conclusion, the flADex trial will address critical research gaps by investigating the acute effects of resistance exercise on CBF, blood biomarkers and cognitive function, areas where evidence remains limited compared to aerobic exercise. This study will provide novel insights into how different exercise modalities acutely influence brain health and neuroprotection during aging.

## Data Availability

All data produced in the present study will be available upon reasonable request to the authors

https://github.com/fladexprojectugr

## ACKNOWLEDGEMENTS

We thank Dr. Veronica Cabanas-Sanchez for performing the randomization of the trial.

