## Supplementary material for "Understanding cerebral blood flow dynamics for Alzheimer’s disease prevention through acute exercise (flADex): Protocol for a randomized crossover trial": flADex_protocol_preprint_MedRxiv_suppl_mod2.docx

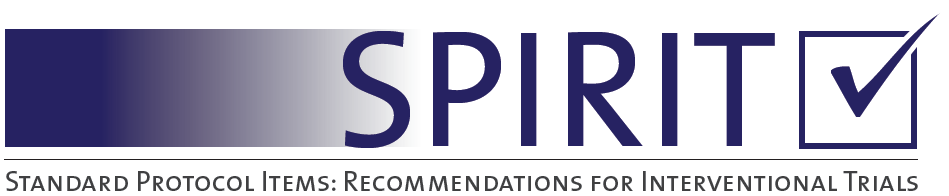

### Supplementary material

#### Table A1

Supplementary table A1. SPIRIT Cheklist of the flADex Trial.

SPIRIT-Outcomes (combination of the SPIRIT 2013 checklist and the 2022 extension) *

| Section/item | Item_No | Description | Addressed headings; pags. |
| --- | --- | --- | --- |
| **Administrative information** | | |  |
| Title | 1 | Descriptive title identifying the study design, population, interventions, and, if applicable, trial acronym | Title page; 0 |
| Trial registration | 2a | Trial identifier and registry name. If not yet registered, name of intended registry | Trial design and ethics; 7 |
|  | 2b | All items from the World Health Organization Trial Registration Data Set | N/A |
| Protocol version | 3 | Date and version identifier | Trial design and ethics; 7 |
| Funding | 4 | Sources and types of financial, material, and other support | Funding; 20, 21 |
| Roles and responsibilities | 5a | Names, affiliations, and roles of protocol contributors | 0 |
|  | 5b | Name and contact information for the trial sponsor | N/A |
|  | 5c | Role of study sponsor and funders, if any, in study design; collection, management, analysis, and interpretation of data; writing of the report; and the decision to submit the report for publication, including whether they will have ultimate authority over any of these activities | Front pag;1 |
|  | 5d | Composition, roles, and responsibilities of the coordinating centre, steering committee, endpoint adjudication committee, data management team, and other individuals or groups overseeing the trial, if applicable (see Item 21a for data monitoring committee) | N/A |
| Introduction |  |  |  |
| Background and rationale | 6a | Description of research question and justification for undertaking the trial, including summary of relevant studies (published and unpublished) examining benefits and harms for each intervention | Background; 4, 5 |
|  | 6b | Explanation for choice of comparators | Overview of experimental condition; 9-11 |
| Objectives | 7 | Specific objectives or hypotheses | Trial objectives; 5 |
| Trial design | 8 | Description of trial design including type of trial (eg, parallel group, crossover, factorial, single group), allocation ratio, and framework (eg, superiority, equivalence, noninferiority, exploratory) | Trial design and ethics; 6.  Figure 1 |
| Methods: Participants, interventions, and outcomes | | |  |
| Study setting | 9 | Description of study settings (eg, community clinic, academic hospital) and list of countries where data will be collected. Reference to where list of study sites can be obtained | Trial design and ethics; 6, 7 |
| Eligibility criteria | 10 | Inclusion and exclusion criteria for participants. If applicable, eligibility criteria for study centres and individuals who will perform the interventions (eg, surgeons, psychotherapists) | Eligibility criteria, screening of participants and recruitment; 7, 8.  Table 1 |
| Interventions | 11a | Interventions for each group with sufficient detail to allow replication, including how and when they will be administered | Overview of experimental conditions; 9-11 |
|  | 11b | Criteria for discontinuing or modifying allocated interventions for a given trial participant (eg, drug dose change in response to harms, participant request, or improving/worsening disease) | Safety and adverse events; 15, 16 |
|  | 11c | Strategies to improve adherence to intervention protocols, and any procedures for monitoring adherence (eg, drug tablet return, laboratory tests) | Resistance exercise condition; 10 |
|  | 11d | Relevant concomitant care and interventions that are permitted or prohibited during the trial | N/A |
| Outcomes | 12 | Primary, secondary, and other outcomes, including the specific measurement variable (eg, systolic blood pressure), analysis metric (eg, change from baseline, final value, time to event), method of aggregation (eg, median, proportion), and time point for each outcome. Explanation of the clinical relevance of chosen efficacy and harm outcomes is strongly recommended | Outcomes; 11-15.  Tables 2, 3 |
| Participant timeline | 13 | Time schedule of enrolment, interventions (including any run-ins and washouts), assessments, and visits for participants. A schematic diagram is highly recommended (see Figure) | Eligibility criteria, screening of participants and recruitment; 7, 8 / Overview of experimental conditions; 9.  Figures 2; 3 |
| Sample size | 14 | Estimated number of participants needed to achieve study objectives and how it was determined, including clinical and statistical assumptions supporting any sample size calculations | Sample size; 16 |
| Recruitment | 15 | Strategies for achieving adequate participant enrolment to reach target sample size | Eligibility criteria, screening of participants and recruitment; 7.  Figure 2 |
| **Methods: Assignment of interventions (for controlled trials)** | | |  |
| Allocation: |  |  |  |
| Sequence generation | 16a | Method of generating the allocation sequence (eg, computer-generated random numbers), and list of any factors for stratification. To reduce predictability of a random sequence, details of any planned restriction (eg, blocking) should be provided in a separate document that is unavailable to those who enrol participants or assign interventions | Randomization; 8, 9 |
| Allocation concealment mechanism | 16b | Mechanism of implementing the allocation sequence (eg, central telephone; sequentially numbered, opaque, sealed envelopes), describing any steps to conceal the sequence until interventions are assigned | Randomization; 8, 9 |
| Implementation | 16c | Who will generate the allocation sequence, who will enrol participants, and who will assign participants to interventions | Randomization; 8, 9 |
| Blinding (masking) | 17a | Who will be blinded after assignment to interventions (eg, trial participants, care providers, outcome assessors, data analysts), and how | Blinding; 9 |
|  | 17b | If blinded, circumstances under which unblinding is permissible, and procedure for revealing a participant’s allocated intervention during the trial | Blinding; 9 |
| **Methods: Data collection, management, and analysis** | | |  |
| Data collection methods | 18a | Plans for assessment and collection of outcome, baseline, and other trial data, including any related processes to promote data quality (eg, duplicate measurements, training of assessors) and a description of study instruments (eg, questionnaires, laboratory tests) along with their reliability and validity, if known. Reference to where data collection forms can be found, if not in the protocol | Outcomes; 11-15.  Tables 2, 3 |
|  | 18b | Plans to promote participant retention and complete follow-up, including list of any outcome data to be collected for participants who discontinue or deviate from intervention protocols | N/A |
| Data management | 19 | Plans for data entry, coding, security, and storage, including any related processes to promote data quality (eg, double data entry; range checks for data values). Reference to where details of data management procedures can be found, if not in the protocol | Data management and sharing; 17, 18 |
| Statistical methods | 20a | Statistical methods for analysing primary and secondary outcomes. Reference to where other details of the statistical analysis plan can be found, if not in the protocol | Data analysis plan; 16, 17 |
|  | 20b | Methods for any additional analyses (eg, subgroup and adjusted analyses) | Data analysis plan; 16, 17 |
|  | 20c | Definition of analysis population relating to protocol non-adherence (eg, as randomised analysis), and any statistical methods to handle missing data (eg, multiple imputation) | Data analysis plan; 16, 17 |
| **Methods: Monitoring** | | |  |
| Data monitoring | 21a | Composition of data monitoring committee (DMC); summary of its role and reporting structure; statement of whether it is independent from the sponsor and competing interests; and reference to where further details about its charter can be found, if not in the protocol. Alternatively, an explanation of why a DMC is not needed | N/A |
|  | 21b | Description of any interim analyses and stopping guidelines, including who will have access to these interim results and make the final decision to terminate the trial | N/A |
| Harms | 22 | Plans for collecting, assessing, reporting, and managing solicited and spontaneously reported adverse events and other unintended effects of trial interventions or trial conduct | Safety and adverse events; 15, 16 |
| Auditing | 23 | Frequency and procedures for auditing trial conduct, if any, and whether the process will be independent from investigators and the sponsor | N/A |
| Ethics and dissemination | | |  |
| Research ethics approval | 24 | Plans for seeking research ethics committee/institutional review board (REC/IRB) approval | Trial design and ethics; 6, 7 |
| Protocol amendments | 25 | Plans for communicating important protocol modifications (eg, changes to eligibility criteria, outcomes, analyses) to relevant parties (eg, investigators, REC/IRBs, trial participants, trial registries, journals, regulators) | Trial design and ethics; 6, 7 |
| Consent or assent | 26a | Who will obtain informed consent or assent from potential trial participants or authorised surrogates, and how (see Item 32) | Trial design and ethics; 6, 7 |
|  | 26b | Additional consent provisions for collection and use of participant data and biological specimens in ancillary studies, if applicable | Trial design and ethics; 6, 7 |
| Confidentiality | 27 | How personal information about potential and enrolled participants will be collected, shared, and maintained in order to protect confidentiality before, during, and after the trial | Data management and sharing; 17, 18 |
| Declaration of interests | 28 | Financial and other competing interests for principal investigators for the overall trial and each study site | Declarations of interest; 2 |
| Access to data | 29 | Statement of who will have access to the final trial dataset, and disclosure of contractual agreements that limit such access for investigators | Data management and sharing; 17, 18 |
| Ancillary and post-trial care | 30 | Provisions, if any, for ancillary and post-trial care, and for compensation to those who suffer harm from trial participation | Safety and adverse events; 15, 16 |
| Dissemination policy | 31a | Plans for investigators and sponsor to communicate trial results to participants, healthcare professionals, the public, and other relevant groups (eg, via publication, reporting in results databases, or other data sharing arrangements), including any publication restrictions | Data management and sharing; 17, 18 |
|  | 31b | Authorship eligibility guidelines and any intended use of professional writers | Data management and sharing; 17, 18 |
|  | 31c | Plans, if any, for granting public access to the full protocol, participant-level dataset, and statistical code | Data management and sharing; 17, 18 |
| Appendices |  |  |  |
| Informed consent materials | 32 | Model consent form and other related documentation given to participants and authorised surrogates | - |
| Biological specimens | 33 | Plans for collection, laboratory evaluation, and storage of biological specimens for genetic or molecular analysis in the current trial and for future use in ancillary studies, if applicable | Secondary outcomes; 13 |

*It is strongly recommended that this checklist be read in conjunction with the SPIRIT 2013 Explanation & Elaboration for important clarification on the items. Amendments to the protocol should be tracked and dated. The SPIRIT checklist is copyrighted by the SPIRIT Group under the Creative Commons “Attribution-NonCommercial-NoDerivs 3.0 Unported” license.

**Supplementary material A2. Exercise conditions**

Below are descriptions of the exercises to be implemented in the flADex trial. All training conditions will be administered to participants in a randomized order.

Graphical presentation of the exercises can be found on the extended version of the supplementary material (<https://github.com/fladexprojectugr>).

**A2.1. Resistance exercise condition**

The resistance exercise condition will last 30 minutes and contains a combination of upper and lower body exercises using elastic bands and body weight exercise of the major muscle groups. The RPE target will be of 4-6 for a moderate intensity. The exercises will be divided into two levels and the level chosen will depend on the physical condition of the participants.

| **Exercise** | **Target muscle** | **Instruction** |
| --- | --- | --- |
| **Cat-camel** | Lumbar, thoracic, and cervical muscles | **Starting position:** Begin standing with hands on the wall, forming a 90º angle, and make sure to stand straight.  **Execution:** Move from a position of lumbar, thoracic, and cervical neutrality to maximum positions of flexion (kyphosis and retroversion), raising the spine. Continue with the extension (lordosis and anteversion), moving the spine downward. |
| **Thoracic mobility** | Extensor of the vertebral column | **Starting position:** Begin standing with one leg forward. Keep the arms extended, facing forward. One hand remains fixed (the same as the forward leg), while the other moves to the opposite side, opening the chest.  **Execution:** Rotate the thorax as much as possible, keeping the body straight. |
| **Hip and knee mobility** | Hip flexors and extensors, dorsiflexors and plantarflexors. | **Starting position:** Stand up. Hands are placed on the hips, and the foot you want to mobilize is positioned forward over the other, pointing towards the wall.  **Execution:** Flex the knee and ankle as much as possible, trying to touch the wall with the knee. Move the forward foot backward to increase the difficulty. |
| **Shouder and hip mobility** | Shoulder, spinal and hip flexors and extensors | **Starting position:** Standing position.  **Elastic band position:** The elastic band is stretched to the maximum without exerting tension with straight arms.  **Execution:** Move the straight arms from the hips to above the head. Abducting the arms to generate tension in the elastic band. When the arms are above the head, perform a squat. |

**Exercise session (26 minutes)**

| **Exercise** | **Target muscle** | **Instruction** |
| --- | --- | --- |
| **Glute bridge for hamstring** | Hamstring and gluteus | **Starting position**: Supine position with bent knees and feet at hip-width, heels on the mat and toes pointing at the ceiling.  **Execution:** Move the hip towards the ceiling, keep the spine neutral and the scapulas flat against the floor. Lift the hips as high as possible. Then, lower the hip in a controlled manner to the starting position.  **Key points:** Perform pelvic retroversion and squeeze the gluteus muscles when hip is high. Push the feet and hands against the floor. |
| **Front plank** | Lumbopelvic musculature | **Starting position:** Prone position with the forearms on the mat supporting the body and elbows at 90º. The knees and feet are at hip- width.  **Execution:** Raise the hip and knees until the trunk is parallel to the mat. Hold the position for 10 seconds and return to the starting position.  **Key points:** Contract the gluteus muscles and abdomen, keep the lumbopelvic neutrality and exhale during the concentric phase. |
| **Standing face pull *** | Trapezius | **Starting position:** Standing position with the feet at hip-width and grab the elastic band with straight arms in pronation with maximum elongation but without tension.  **Elastic band position:** Nose height.  **Execution:** Pull the elbows towards the face and bend the elbows to 90º. Keep the elbows on shoulder-height.  **Key points:** Keep the elbows at shoulder-height, and the shoulders down. |
| **Incline push up** | Pectoral | **Starting position:** Standing position one meter in front of the object (e.g., a table). Place the hands on the edge of the table slightly wider than shoulder-width. Arms are straight.  **Execution:** Bend the arms, so that the chest approaches the object as much as possible. Hold for a second, extend the arms and return to starting position.  **Key points:** Maintain a plank position, push with the hands, and align the gluteus muscles and the trunk. |
| **Squat** | Quadriceps and gluteus | **Starting position:** Standing position with the legs hip-width.  **Execution:** Lower into a squatting position and keep the lumbar in a neutral position. During this movement, the hip, knee and ankle are bent. After this, extend the legs and return to starting position.  **Key points:** Push with the legs to get up and keep the knees aligned with the legs during the whole exercise. |
| **Press pallof *** | Lumbopelvic musculature | **Starting position:** Standing position with the elbows bent at 90º and the feet at hip-width. Grab the elastic band on the side at the maximum elongation without tension.  **Elastic band position:** Chest level.  **Execution:** Exhale, stretch the arms and hold for 3 seconds. Return to the starting position while inhaling. Focus on breathing when stretching the arms. Perform 20” continuously to each side.  **Key points:** Control the breathing, move slowly, and controlled during the eccentric phase**.** |

| **Lunge** | Quadriceps and gluteus | **Starting position:** Standing position, with one leg in front of the other and with approximately 0.6m.  Back leg stands on the toes and the hands are placed on the hips.  **Execution:** Bend both knees to 90º. Stand back up and lift the back foot from the ground. Swing the back foot forward and step forward.  **Key points:** Align the front knee and leg, push the foot against the floor, and push with the front leg. |
| --- | --- | --- |
| **Seated shoulder press *** | Deltoid | **Starting position:** Sitting position and pass the elastic band under the seat of the chair. Grab the elastic band neutrally.  **Elastic band position:** Underneath the seat of the chair.  **Execution:** Push the arms above the head until the arms are straight. Align the wrists with the arm during the movement.  **Key points:** Push from the feet to the shoulder. |
| **Glute bridge for hamstring** | Hamstring and gluteus | **Starting position**: Supine position with bent knees and feet at hip-width, heels on the mat and toes pointing at the ceiling.  **Execution:** Move the hip towards the ceiling, keep the spine neutral and the scapulas flat against the floor. Lift the hips as high as possible. Then, lower the hip in a controlled manner to the starting position.  **Key points:** Perform pelvic retroversion and squeeze the gluteus muscles when hip is high. Push the feet and hands against the floor. |
| **Kneeling plank** | Lumbopelvic musculature | **Starting position:** Prone position with the forearms supported on the mat. The knees and feet are at hip-width on the mat.  **Execution:** Raise the hip and keep the knees and feet on the mat. The trunk is parallel position with the mat and the elbows are bent at 90º. Hold the position for 10 seconds and return to the starting position.  **Key points:** Strengthen the gluteus muscles and core and maintain the lumbopelvic in a neutral position. Exhale during the concentric phase. |
| **Standing face pull *** | Trapezious | **Starting position:** Standing position with the feet at hip-width and arms straight. Grab the elastic band in pronation with maximum elongation but without tension.  **Elastic band position:** Nose height.  **Execution:** Pull the elbows towards the face and bend the elbows at 90º. Keep the elbows on shoulder-height.  **Key points:** Keep the elbows at shoulder-height and the shoulders down. |
| **Wall push up** | Pectoral | **Starting position:** Standing position one meter in front of the wall. The hands are placed at the wall on shoulder height and width.  **Execution:** Bend the arms, so that the face approaches the wall as much as possible.  **Key points:** Maintain the plank position, push with the hands, and align the gluteus muscles with the trunk. |
| **Squat with crossed arms** | Quadriceps and gluteus | **Starting position:** Standing position in front of the chair with the legs at hip-width and the hands crossed on the chest.  **Execution:** Lower into a squatting position and lightly touch the chair, then get up from the chair.  **Key points:** Push with the legs to get up from the chair, keep the knees aligned with the legs during the whole exercise. |
| **Press pallof *** | Lumbopelvic musculature | **Starting position:** Standing position with the elbows bent at 90º and the feet at hip-width. Grab the elastic band on the side at the maximum elongation without tension.  **Elastic band position:** Chest level.  **Execution:** Exhale, stretch the arms and hold for 3 seconds. Return to the starting position while inhaling. Focus on breathing when stretching the arms. Perform 20” continuously to each side.  **Key points:** Control the breathing, move slowly, and controlled during the eccentric phase**.** |
| **Lunge** | Quadriceps and gluteus | **Starting position**: Standing position with one foot in front of the other with the distance of approximately 0.6 m. Place the hands on the hips.  **Execution:** Bend both knees to approximately 90°. With this movement, the back knee is hovering just of the floor and the front knee and ankle are aligned. Return to the starting position by extending the knees. Perform 20” continuously with each leg  **Key points:** Align the front knee and the foot and push the feet against the floor. |
| **Seated shoulder press *** | Deltoid | **Starting position:** Sitting position with legs fully supported. Pass the elastic band under the seat of the chair and grab it neutrally.  **Elastic band position:** Under the chair seat.  **Execution:** Push the arms up until elbows are straight. Align the wrists with the arm during the exercise.  **Key points:** Push from the feet to the shoulder. |

**A2.2. Moderate aerobic exercise condition**

The aerobic exercise condition will last 30 minutes of continuous moderate intensity aerobic exercise on a bike. The intensity target will be at 60%-70% of their maximal heart rate (HRmax).

**Warm-up (4 minutes)**

| **Step** | **Instruction** |
| --- | --- |
| **1** | Adjust the saddle height so that your extended leg has a slight bend at the knee while pedaling. |
| **2** | Ensure the saddle is horizontal to prevent discomfort; adjust the angle to make it parallel to the ground. |
| **3** | Adjust the handlebars to a height that allows you to maintain an upright and comfortable posture. |
| **4** | Check the distance between the saddle and the handlebars; it should allow you to reach them without overstretching or being too cramped. |
| **5** | Before starting, verify that all adjustments are secure and tight, avoiding any looseness or unexpected movements |

**Exercise session (26 minutes)**

| **Exercise** | **Target muscle** | **Instruction** |
| --- | --- | --- |
| **Bike** | Quadriceps, hamstrings, glutes, calf, hip muscles, abdominal muscles, and lower back muscles. | A continuous aerobic exercise is performed on a stationary bike, which will be within the at 60%-70% of their maximal heart rate (HRmax). |
